# Social heterogeneity drives complex patterns of the COVID-19 pandemic: insights from a novel Stochastic Heterogeneous Epidemic Model (SHEM)

**DOI:** 10.1101/2020.07.10.20150813

**Authors:** Alexander V. Maltsev, Michael D. Stern

## Abstract

In today’s absence of a vaccine and impactful treatments, the most effective way to combat the virus is to find and implement mitigation strategies. An invaluable resource in this task is numerical modeling that can reveal key factors in COVID-19 pandemic development. On the other hand, it has become evident that regional infection curves of COVID-19 exhibit complex patterns which often differ from curves predicted by forecasting models. The wide variations in attack rate observed among different social strata suggest that this may be due to social heterogeneity not accounted for by regional models. We investigated this hypothesis by developing and using a new Stochastic Heterogeneous Epidemic Model (SHEM) that focuses on subpopulations that are vulnerable in the sense of having an increased likelihood of spreading infection among themselves. We found that the isolation or embedding of vulnerable sub-clusters in a major population hub generated complex stochastic infection patterns which included multiple peaks and growth periods, an extended plateau, a prolonged tail, or a delayed second wave of infection. Embedded vulnerable groups became hotspots that drove infection despite efforts of the main population to socially distance, while isolated groups suffered delayed but intense infection. Amplification of infection by these hotspots facilitated transmission from one urban area to another, causing the epidemic to hopscotch in a stochastic manner to places it would not otherwise reach, resembling a microcosm of the situation worldwide as of September 2020. Our results suggest that social heterogeneity is a key factor in the formation of complex infection propagation patterns. Thus, the mitigation of vulnerable groups is essential to control the COVID-19 pandemic worldwide. The design of our new model allows it to be applied in future studies of real-world scenarios on any scale, limited only by computing memory and the ability to determine the underlying topology and parameters.

## Introduction

Coronaviruses represent one of the major pathogens that primarily target the human respiratory system. Previous outbreaks of coronaviruses (CoVs) that affected humans include the severe acute respiratory syndrome (SARS)-CoV and the Middle East respiratory syndrome (MERS)-CoV [1]. COVID-19 is a disease caused by the novel coronavirus SARS-CoV-2 virus that is both fatal and has a high transmission rate (*R*_*0*_), almost twice that of the 2017-2018 common influenza [2, 3]. The World Health Organization stated that this combination of high health risk and susceptibility is of great global public health concern, and efforts must be directed to prevent further infection while vaccines are still being developed [4]. As of November 2020, there are almost sixty million confirmed COVID-19 cases worldwide and close to confirmed one and a half million deaths. Older adults seem to be at higher risk for developing more serious complications from COVID-19 illness [5, 6]. In today’s absence of a vaccine and impactful treatments, the most effective way to combat the virus is to find and implement mitigation strategies. An invaluable resource in this difficult task is numerical modeling studies that can reveal key factors in pandemic development.

What models could be useful? Direct study of the available data of COVID-19 is complicated because many cases and deaths are underrepresented. However, a simple model that correctly captures large-scale behaviors, but gets some details wrong, is useful, whereas a complicated model that gets some details correct but mischaracterizes the large-scale behaviors is misleading [7]. Previously, during the H1N1 pandemic, generic (i.e. non-specific) stochastic influenza models were important to understand and quantify the full effects of the virus in simulations of important scenarios [8]. Open source stochastic models such as FluTE (2010) or GLEaM (2011) [9, 10] were developed to simulate the spatial interaction and clusterization of millions of people to discover epidemic patterns.

Now, with respect to COVID-19, the FluTE model has recently been used to offer interventions to mitigate early spread of SARS-CoV-2 in Singapore [11], and GLEaM was adopted by Chinazzi et al. [12] to model the international propagation of COVID-19 to gain insight into the effect of travel restrictions on virus spread. Detailed statistical information about the social interactions and grouping of individuals is difficult to gather, but ultimately can be used to calibrate the parameters of agent-based models. Such calibrated agent-based models have been applied to model high-density housing in Brazil and their effect on viral spread to the rest of the population [13].

Despite extensive efforts to understand and predict the COVID-19 spread, the key factors that determine the multimodal rise patterns, the asymmetry of the recovery phase, and the emergence of a distinct second wave remain unclear. Therefore, instead of another data-based forecasting model, we chose to develop a scenario model to study the consequences of a set of hypothesis-driven conditions in a network of populations. One underexplored but important factor of pandemic spread is social heterogeneity which defines the degree of dissimilarity in the behaviors of embedded subpopulations. With regard to virus spread, the important characteristics of social heterogeneity to consider are levels of clusterization, societal interaction, and disease mitigation strategies. Our hypothesis is that complex infection curves that consist of multiple infection peaks and growth periods are the consequence of asynchronous propagation of infection among groups with widely varying degrees of intra-group interaction and isolation from main hubs (a metapopulation of infections).

To approach this problem, we developed a novel Stochastic Heterogeneous Epidemic Model (dubbed SHEM) which incorporates heterogeneous aspects of society. We also take into account over-dispersed stochasticity (super-spreading) [14], which is usually not incorporated into compartmental models but can be critical in small or virgin populations. The model design was inspired by our stochastic models of local calcium release dynamics inside heart cells, driven by explosive calcium-induced-calcium-release [15, 16]. We examine several key scenarios of heterogeneity where separate communities of various clusterization and transmission capabilities are linked to a large population hub. The basic reproduction number of infection (*R*_*0*_) of the bulk of our population was assigned to *R*_*0*_ = 2.5 which is within the range of SARS-CoV-2 basic reproduction number based on the early phase of COVID-19 outbreak in Italy [17]. Interplay of various degrees of heterogeneity and isolation periods in our model generated various dynamic patterns of infection, including a multi-modal growth periods, an extended plateau, prolonged tail, or a delayed second wave of infection. Most importantly, we found that vulnerable social subgroups play a key role in the propagation and unpredictability of the epidemic, and can defeat efforts at social distancing.

## Methods

### Model purpose

In view of the constantly changing behavioral environment for COVID-19 in the United States and worldwide, data-based predictive modeling of the future of the epidemic is difficult. Our model is specifically intended to examine the effect of heterogeneity, including not only geographic but also social heterogeneity, i.e. the existence of groups within one geographic location that have different social interaction patterns and may be partially isolated from neighboring groups, *e*.*g*. nursing homes, prisons, campuses. Alternatively, subgroups can be partially embedded in the main population, e.g. meat processing plants or warehouse employees who are unable to socially distance at work, but spend part of their day in the main community where they can acquire and amplify infection. The model is fully stochastic and, unlike most compartmental models, incorporates the effect of over dispersion of secondary infections (super spreading).

### Structure of the Model

The general model consists of a number of subpopulations (“villages”) whose number is limited only by computing memory. The simulation is based on a generalization of the SEIRD representation. The state of each village is represented by the numbers of individuals in each of 5 states: *Susceptible, Exposed* (destined to become infected), *Infected, Recovered* (immune) and *Dead* (however, see below under Super-spreading for additional state-dependence). Each village is, by definition, homogeneous and mixed. Villages could represent actual geographic units, but could also be groups or sub-regions that have different social interactions or behavior. The mean duration of infection (infectious period) was taken to be 7 days and the incubation period 5.5 days.

Each village J is characterized by its population, the expected mortality of virus infections, and its local value RINN(J) of the basic reproduction number *R*_*0*_. *R*_*0*_ is defined as the mean expected number of secondary infections spawned by one infected individual over the duration of their illness, *if the population were totally susceptible*. It is a property of both the virus and the behavior of individuals in the population, but is distinct from *R(t)*, the realized, time dependent, reproduction number that depends also on the fraction of susceptible individuals remaining during the epidemic.

Villages are connected by a user-specified network of formally unidirectional links along which infection or individuals can travel at user-specified rates, including links from each village to itself to represent internal infection/recovery processes. Infection can spread by two processes: transient contact between groups (*alpha* process) *e*.*g*. nursing home staff coming from the city; or actual migration of individuals from one village to another (*beta* process). Each non-self link is characterized by 4 user-supplied parameters: *alphain* and *alphaout* describe the degree of transient contact (see below) along or against the direction of the link respectively; *betain* and *betaout* are rates of migration of individuals (time^-1^).

### Transient Contact (alpha) Process

Infection transmitted by transient contact is modeled as though members of one village spend some (small) fraction *alpha(in/out)* of their time (*i*.*e*. of their inter-personal contacts) “visiting” the opposite village at the other end of the link, adjusted for any mitigations (an example would be staff working at a nursing home, or meat-packing plant employees, treated as a separate, high-risk population but living in the surrounding county). The spread of infection in each direction of the link has two components: (1) exposure of susceptibles by visiting infectious individuals and (2) exposure of visiting susceptibles in the visited village, who then carry the infection back to their village. This formulation allows for the possibility that transmission is asymmetric. The generation of exposure by these “visitors” at home and abroad is scaled so that each infected individual, generates (in an otherwise susceptible population) his destined number of secondary cases (see below under super-spreading).

This arrangement allows for the possibility that “visitors” from different villages could cross-infect while visiting a common hub (picture UPS and FEDEX drivers) even if there is no direct link between them. To represent this process, “virtual links” are generated between pairs of physical links that meet in a hub (in graph-theory terms these are links of the adjoint graph of the network). Infection by this indirect process is second-order in the alpha’s so it makes very little contribution in the case of highly isolated sub-populations (*e*.*g*. nursing homes, prisons) but could be important for embedded sub-populations with high contact with the hub. Although each village is considered homogeneous by definition, further heterogeneity within a village could be represented by subdividing the population into several “villages” in close mutual contact via the alpha process (*e*.*g*. students in a college split into those who go to bars and those who study alone).

### Simulation Method

The entire collection of populations is simulated as a single, continuous-time Markov chain (birth-death process). There are 16 types of possible events associated with each link:

1. Infection from source to target by transient contact
2. Infection from target to source by transient contact
3. Infected individual moves from source to target
4. Exposed individual moves from source to target
5. Susceptible individual moves from source to target
6. Infected individual moves from target to source
7. Exposed individual moves from target to source
8. Susceptible individual moves from target to source
9. Susceptible gets exposed inside village (self-link only)
10. Exposed converts to infected inside village (self-link only)
11. Infected recovers inside village (self-link only)
12. Infected dies inside village (self-link only)
13. Recovered moves from source to target
14. Recovered moves from target to source
15. Susceptible gets vaccinated
16. Recovered loses immunity

The objective of the simulation is to generate a continuous-time sequence of Markov states, with transition rates determined by the SEIRD equations, modified as described below under Super Spreading. The algorithm consist of a front-end program that sets up the network of villages and the rates of spread of infections by the alpha and beta processes, and an engine module that is called repeatedly by the front-end to walk the Markov scheme under a sequence of imposed conditions, *e*.*g*. open, lockdown etc. The operation of the program is described by the following simplified pseudocode:

~~~
PROGRAM FRONT_END
use module simulator
read parameter file !nh=number of villages
               do ih=1,nh
               initialize village population sizes and states
               lrlinks(ilink,1:2)=ih !create self-links
               set r0’s for first time period
               end do
!create network
        lrlinks(ilink,1)=source
        lrlinks(ilink,2)=target
        set alphain,alphaout,betain,betaout(ilink)
        ilink++
call episim(…lrlinks…tswitch,yflag) !invoke the engine in simulator module
if yflag=false on return then ! t reached a breakpoint
        change r0’s,alphas,betas
        advance tswitch
        call episim again
else
        reached tmax
        write output history
end program front-end
MODULE SIMULATOR
contains
subroutine episim ! main engine
create bidirectional linked infectivity lists
!generate virtual links by extending link array
do ll1=1,nrlinks
        do ll2=ll1,nrlinks ! triangular search for common hubs j3
        ilink++
        links(ilink,3)=j3
        alphav(ilink)=alphain/out(j1)*alphaout/in(j2)
        end do
end do
t=0
! main loop
do while t<tswitch
        do over all links
                do event=1,16 !generate cumulative rates of possible events
                rtot=rtot+rate(event,link)
                rtt(jtt)=rtot
                jtt++
                end do
        end do
! rtot is total rate of available markov transitions
! time of next event in Poissant point process
        time of next event = t-log(random)/rtot !exponential distribution
! choose the actual event link and type:
        find rtot*random2 in the cumulative array rtt at index jbin
        jl=(jbin-1)/16+1 ! find which link fired
        links(jl,1:3) gives the villages at the link ends and/or hub
        jp=jbin-16*(jl-1) ! remainder points to the event type
! carry out the event
        if the event creates a new infectious person then
                k=kranbin(random3,rinn(j),reff) ! personal infectivity
                push k on the top of infection list of village j
                inf(j)=inf(j)+k
! inf is the collective infectiousness of village j, plays role of
! (numberinfected)*r0 in SEIRD equations
        end if
        if the event removes infectious person by recovery or death then
                pull k off bottom of infection list
                inf(j)=inf(j)-k
        end if
        if the event is migration of infectious move between tops(most recent) of
                infection list
        end if
        if t>tmax then
                return with yflag=true
        end if
        if t>tout then
                record state in kout array
                increment tout
        end if
        end do over links
end do while ! continue with time steps until t>tswitch
        return with yflag=false ! continue to the next simulation period.
end subroutine episim
FUNCTION kranbin ! draws random negative binomial integer with mean r0 and          !
dispersion reff.
end module simulator
~~~

### Super-spreading

It is known that the distribution of secondary COVID-19 infections generated by a single, infected individual is over-dispersed (*i*.*e*. has a long tail compared to the Poisson distribution of infections expected if transmission were random). Although the average *R*_*0*_ is estimated to be 2.5-4 in the absence of social distancing mitigations, contact tracing has shown that single individuals have infected up to a hundred others. This is known as super-spreading events, and can occur by several possible mechanisms, involving either a predilection of an individual (*e*.*g*. a celebrity who travels widely and contacts many other people) or a situation in which individuals were placed in unusually close contact (*e*.*g*. a church choir in an indoor location). On the other hand, the majority of infected individuals do not appear to spread the infection to anyone. It has been shown [14] that this over-dispersed distribution can be approximated by a negative binomial distribution, with mean *R*_*0*_ (by definition) and dispersion parameter *r<<1*, for example 3 and 0.16. By iterating this distribution for several generations of viral spread, it is found that the eventual distribution of epidemic size is predicted to be quite different than found for a hypothetical stochastic transmission by Poison-distributed secondary infections with the same *R*_*0*_. A recent model of contact tracing assumed, based on data from the Netherlands, that the distribution of number of personal contacts outside the family is distributed as a negative binomial and used this to generate random changes to infection levels at 1-day intervals [18].

Unfortunately, viral generations do not remain synchronous in time, so it is not straightforward to incorporate super-spreading in a time-dependent epidemic evolution model except by following the interactions and infections of each individual in the population, as done for example in the FLuTE simulation for influenza [8]. This is very compute-intensive, but a more significant objection from our point of view is that it depends on knowing (statistically) the social interaction groups and travel behavior of the population at a fine-grained scale, and these have been severely disrupted by mitigation efforts during the current pandemic. It is possible to try to adjust for these mitigations by calibration against the evolving case data, but this is difficult. Rather than speculate on these variables, we have developed a modified Markov scheme that tries to reproduce the observed distribution of secondary infections by replacing *R*_*0*_ in the event-rate calculations by an infectivity that is itself stochastic. This requires storing a partial history of individual infections, which makes the actual state-space, considered as a Markov process, much larger than that in a classic SEIRD model.

The stochastic process of infection generation by one infected case is in competition with the independent stochastic recovery process. In the model, recovery is a Poisson point process with a rate proportional to the number of infections. If we don’t identify individuals, a super-spreader is likely to be “recovered” before (or after) generating his destined number of infections. To avoid this, we have adopted the following scheme:

- In each village *j*, at each event, an infectivity *inf(j)* is maintained that takes the place of *k*_*i*_*\*R*_*0*_ in the SEIRD rate equations.
- Whenever a new infection is created (by conversion of an exposed individual), a random number *K* is drawn from a negative binomial distribution of mean *R*_*0*_ and dispersion *r*_*eff*_,
- the latter to be determined. *Inf* is incremented by *K* and the individual infectivity *K* is placed on the top of a linked list.
- Whenever a random recovery event is generated at the above-mentioned rate, the oldest individual infectivity is removed from the bottom of the list and subtracted from *inf*.

. The number of secondary infections actually realized by one infected individual is proportional to the actual length of time he remains infectious. Since infections recover in the order in which they were created, if there are *n* infections active, that lifetime will be the *n*^th^ waiting time of the Poisson point process whose rate is *n* times the mean recovery rate (*i*.*e*. the reciprocal of the mean infection duration). The secondary infections generated by individual *K* are a Poisson point process, which is then convolved with the recovery process to give the realized distribution of secondary infections generated by that individual. Further convolving that with the negative binomial distribution of *K* with mean *r*_*0*_ and dispersion *r* we find:

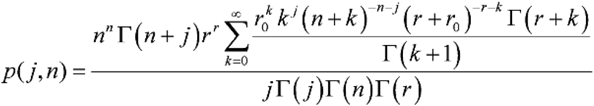

as the distribution of the actual, realized number of secondary infections. This is a long-tailed probability distribution that can be fit, by an appropriate choice *r*_*eff*_ for the dispersion parameter *r* so as to approximate the empirical negative binomial distribution with *r=0*.*16* over the relevant range. With more than a few active infections present, the distribution converges to the limit:

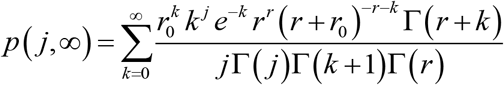

We choose *r*_*eff*_ to give the best least-squares fit on a linear scale for the case *n=1*, which is the most important stochastic case since it governs the chance that a single infected individual can start an outbreak, and gives the chance that an infected individual causes no secondary infections, *p(0,1)=0*.*62* similar to the empirical distribution. These distributions are all normalized and have mean *R*_*0*_ and differ dramatically from the Poisson distribution (Fig S3, dashed line) assumed in the classic SEIR model. Larger values of *n* are decreasingly important because the aggregate distribution of the actual infection rate controlled by the sum *inf* behaves similarly to *negbinomial (R*_*0*_,*n*r)* which converges to Poisson, so stochastic effects become less important once there are many active cases.

### Super-spreaders vs super-spreading events

Super-spreading can be a property of the individual or of the circumstances. What happens when an individual infected patient migrates to a new village? Does he keep his identity or does he assume the infectiousness typical of the local *R*_*0*_ of his new environment? In the model we can make the choice, determined by a logical variable SPREADR (default TRUE. controlled in the demos by the input parameter *spreads*). If SPREADR is true, a migrant keeps his prior *K* value which simply migrates from the top (newest) link to be added to the top of the infection list in the new village, thereby preserving his infectious lifetime in his new home. If *SPREADR* is false then the *K* value of migrants is re-randomized using the local *R*_*0*_ and *r*_*eff*_ and the infectivity of transient visitors in the alpha process is re-scaled to the local value of *R*_*0*_. In the current version of the program, SPREADR is a single variable governing all events, but it could easily be made specific to individual links to distinguish groups that are vulnerable due to high density in their home village (e.g. factory or warehouse) versus groups that are intrinsically super-spreaders due to their individual behavior (celebrities, bar hoppers).

### Software Considerations

The model software is written in Fortran 77/95. The main simulation engine, described above, is in the form of a single Fortran module SIMULATOR. It is intended to be driven by a front-end program that sets up the network and scenario. For purpose of these demonstrations, we hand-coded a front end (epichainF) describing a chain of urban clusters (or a single cluster) connected by bidirectional travel, each linked to a large set of small subpopulations whose characteristics differ from the urban cluster. The single Markov-chain structure of the model is intrinsically serial, and is implemented in a single processor thread. For networks with many nodes and dense links this can be speeded up about 5-fold with 32 processors by parallelizing an inner loop.

## Results

### Simulations of infection in isolated clusters driven by an urban cluster

In the first set of simulations we examined the virus spread in simple hypothetical scenarios with equal numbers of individuals in urban and isolated populations (Fig 1A, insert). The large urban cluster was composed of 1 million individuals set to *R*_*0*_=2.5 (open level, but changing throughout the simulation). The isolated population consisted of 250 clusters, each with 4000 +/- 500 people and with the same internal *R*_*0*_=2.5 that remained constant throughout all simulation stages. The urban cluster was weakly connected with 0.001% transient contact into the isolated clusters (*alphainpop*) while isolated clusters had 0.1% contact into the urban cluster (*alphaoutpop*), see Methods for the definition of transient contact. This can be visualized as a collection of small suburban neighborhoods or nursing homes that are attempting to isolate themselves from the city. We investigated 4 scenarios, specified below. In each scenario except #1, the urban cluster closed to *R*_*0*_=1.25 at t=40 days (closed level, e.g. this was New York City under lockdown, based on 21% antibody positive tests at the peak [19]).

**Fig 1:**
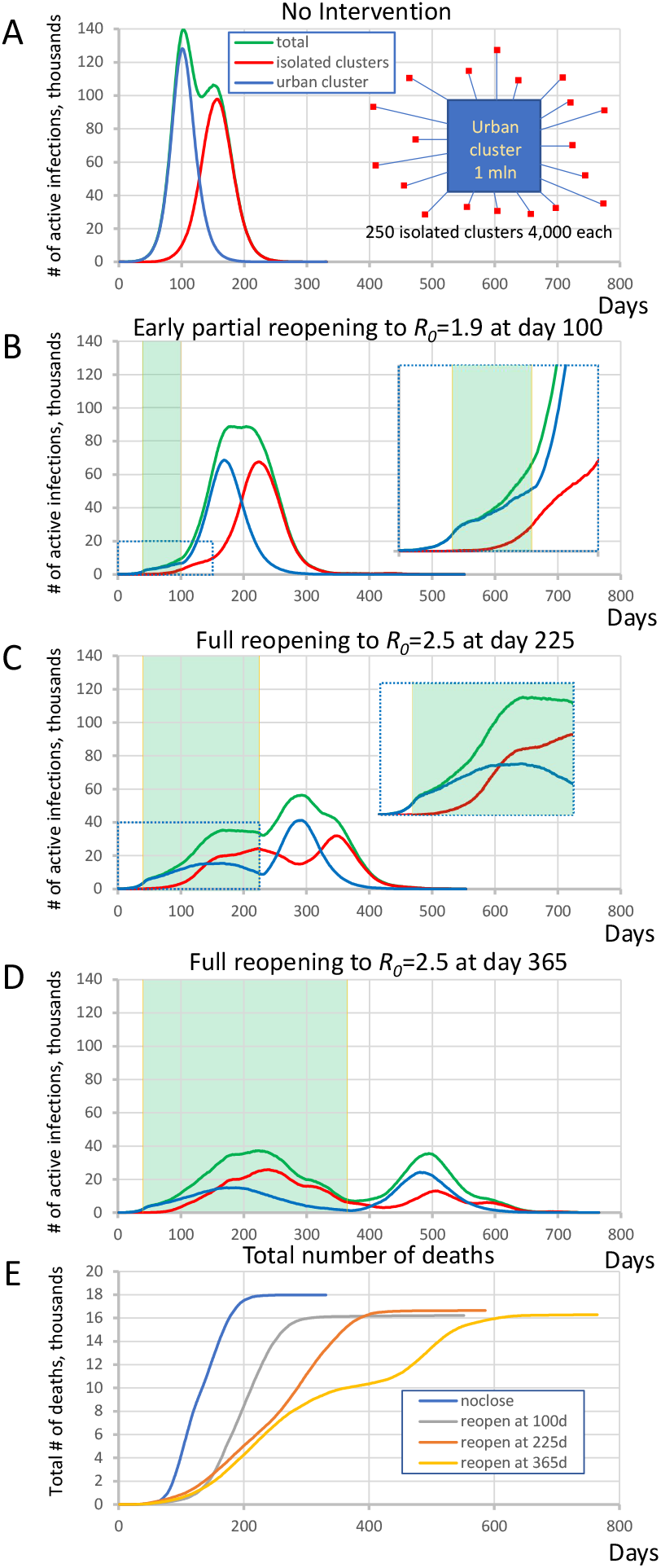
Complex dynamic patterns of SARS-CoV2 infection in simulations in a heterogeneous society when infection in isolated clusters are ignited by an urban cluster implementing various lockdown strategies. (**A)** isolated clusters generate a second delayed peak when no intervention is implemented. Inset schematically illustrates the society structure in this scenario. Contributions are shown by different colors. (**B)** an apparent plateau after early reopening and complex rise pattern during close period (inset). Green shade shows the lockdown periods. (**C)** A multimodal rise (inset) with additional peak generated by rural cluster after full reopening at day 225. (**D)** A delayed second wave emerged after full reopening at day 365. **E**, The dynamics of total number of deaths in each scenario.

1. No mitigation, i.e. freely expending pandemic: The large cluster of individuals stays always open.
2. Premature, partial reopening to *R*_*0*_ = 1.9 at 100 days.
3. Moderate lockdown period with full reopening at 225 days to *R*_*0*_ = 2.5.
4. Long lockdown period with full reopening at 365 days to *R*_*0*_ = 2.5.

A general tendency throughout all 4 scenarios was that as the lockdown period increased, the magnitude of the infection decreased but its duration increased. At the same time, the interplay of the urban cluster and the isolated clusters generated a variety of specific patterns in virus spread dynamics. In the first “no mitigation” scenario (Fig 1A) the isolated areas generated a strong second peak at the time when infection in the urban cluster had gone through its peak and was decaying. On the other hand, the infection rise in the “premature reopening” scenario (Fig 1B) was multi-modal, and the cumulative peak in isolated clusters happened later than the urban cluster, creating an apparent plateau in active infection cases from day 175 to 225. The infection dynamics in the “moderate lockdown” scenario (Fig 1C) was more complex. During the closed stage (of the urban center), the infection in the urban cluster declined, but the delayed infection in isolated clusters continued to rise forming an additional peak in total infections (Fig 1E, inset). Then another peak in total infections emerged in the reopen stage that was generated mainly by the urban cluster, and then was echoed by the isolated subpopulations. Finally, in the “late reopening” (Fig 1D) scenario, infection decreased during the first wave in both urban and isolated clusters but a distinct delayed second wave of infection occurred.

We also performed a control simulation to validate that heterogeneity of isolated clusters is indeed important for the infection pattern. In the most complex scenario of “moderate lockdown” shown in Figure 1C we substitute 250 clusters by one big cluster with the same population of one million people keeping all other parameters the same. The simulations showed a different pattern in which the second big cluster always generated a peak of substantially larger amplitude (Fig S1).

### Simulations of integrated clusters driving infection in an urban cluster

By altering parameters in the same topology as Figure 1A, we found that the outlying clusters, if they are unable to socially distance, can become potential “hotspots” that can drive the infection in the urban population even against efforts of the latter to lock down. In this scenario the large urban cluster was composed of 1 million individuals with *R*_*0*_ = 1.25 throughout all simulation stages while the highly susceptible population consists of 250 clusters each with 1200 +/- 500 people and internal *R*_*0*_ = 3.0 that are partially embedded in the urban cluster. This *R*_*0*_ value is based on data from four districts in Germany when essential manufacturing sectors were open – 95%-prediction interval: 2.16 – 3.73 [20]. The potential hotspot clusters were connected with 20% out-coupling into the urban cluster (*alphaoutpop = 0*.*20*, see Methods). This mechanism of transient contact implements short-term movement of the same people in and out regularly, which does not dilute the effect of the conditions in hotspots the way that random bidirectional migration would. In other words, the same people “virtually” move back and forth but spend most of their time in the high-*R*_*0*_ locations where the infection regenerates. In this scenario, the small number of infections in the urban area are picked up by hotspots, amplified, and then drive a wave of infection among the urban population despite their efforts to keep their internal *R*_*0*_ at 1.25 by social distancing.

We performed 10 runs of these simulations which demonstrated that the integrated clusters drove infection in the urban cluster as shown in a typical example in Fig 2A, B, leading the late appearance of the epidemic in places that had seen few cases in a microcosm of the pattern. In the second “chain” topology multiple small urban areas (population 100K each) are sequentially connected and 30 potential hotspots with *R*_*0*_=2.0 drive infection within each urban cluster and facilitate propagation from cluster to cluster (Fig 3, Fig S2, and Movie S2 show the stochastic dynamics of individual hotspots). In this model, the first urban cluster began with *R*_*0*_ = 2.5, then locked down to 1.25 at day 40, while the unsuspecting urban clusters connected through the chain kept *R*_*0*_ = 1.25 throughout, signifying efforts at social distancing. Ultimately these efforts were defeated by the hotspots picking up the small number of arriving infections and amplifying them. These results demonstrate that subgroups who cannot or will not socially distance can drive the propagation of the epidemic to new regions against the best efforts of the majority of the populations. It follows that it is possible to control the spread of the epidemic through the mitigation of hotspot amplification. To validate this finding, we simulate the application of vaccine treatments to *just* the hotspot members, who constitute only about 30% of the population. The vaccine treatment is applied to individuals in hotspots at the rate of 5% per day, and, as a result, the geographic spread of infection is sufficiently stopped and the entire downstream region is protected from infection and deaths (Fig 4).

**Fig 2:**
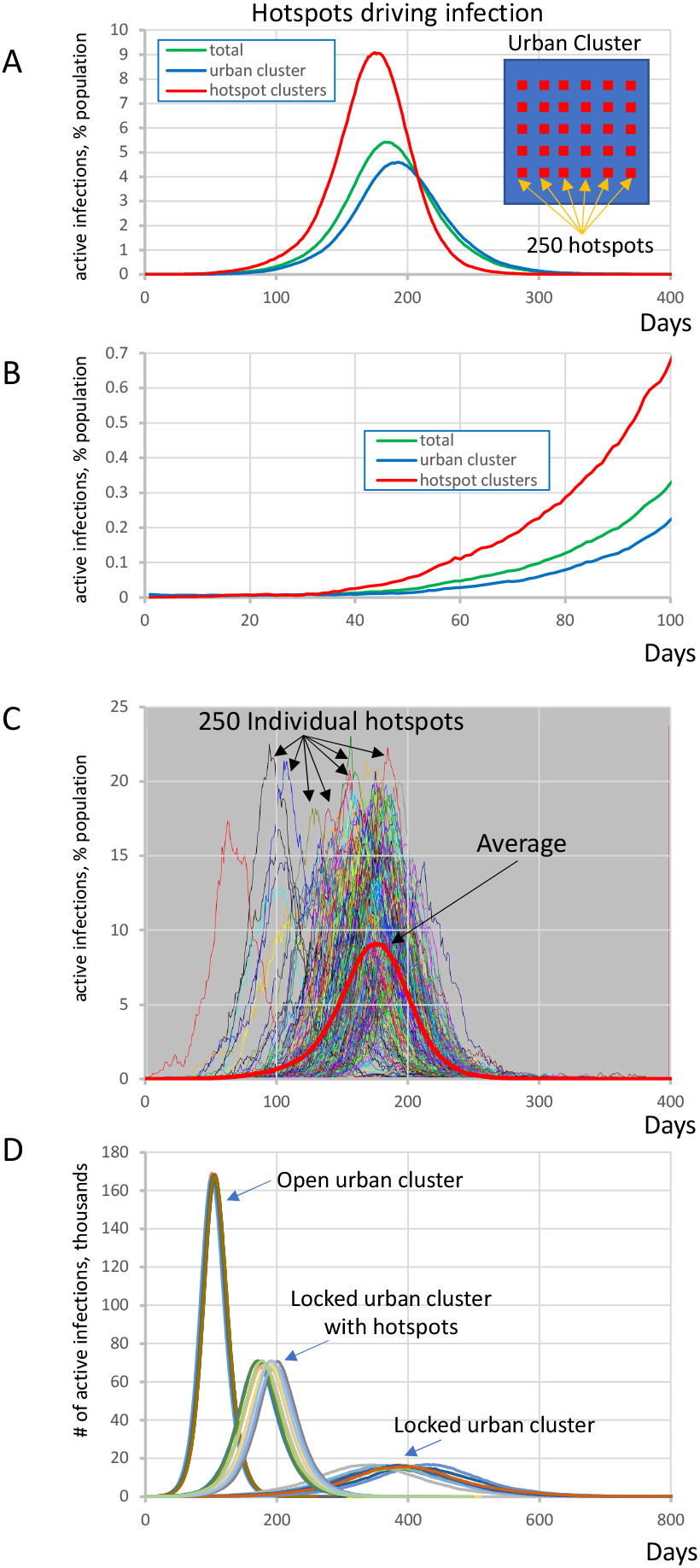
Highly susceptible integrated clusters (hotspots) drive SARS-CoV2 infection in an urban cluster. (**A and B)** Initial rise of infection in hotspot clusters is followed by the infection in urban cluster with a delay of about 30 days. Y-axis represents active infections in % population reflecting for hotspots (red line) the ratio of all active cases in all hotspots to entire population of all 250 hotspots. Inset shows schematically the society structure in this scenario. (**C)** Infection in individual hotspots (multiple colors) substantially fluctuates in terms of time of ignition and magnitude from the mean (red bold curve). See also Movie S1. **(D)** Explosive infection in hotspots within locked urban cluster substantially increased the peak of infection in the entire society and shifted it towards much earlier occurrence from about 400 days to 200 days. Shown are 10 simulation runs for each scenario.

**Fig 3:**
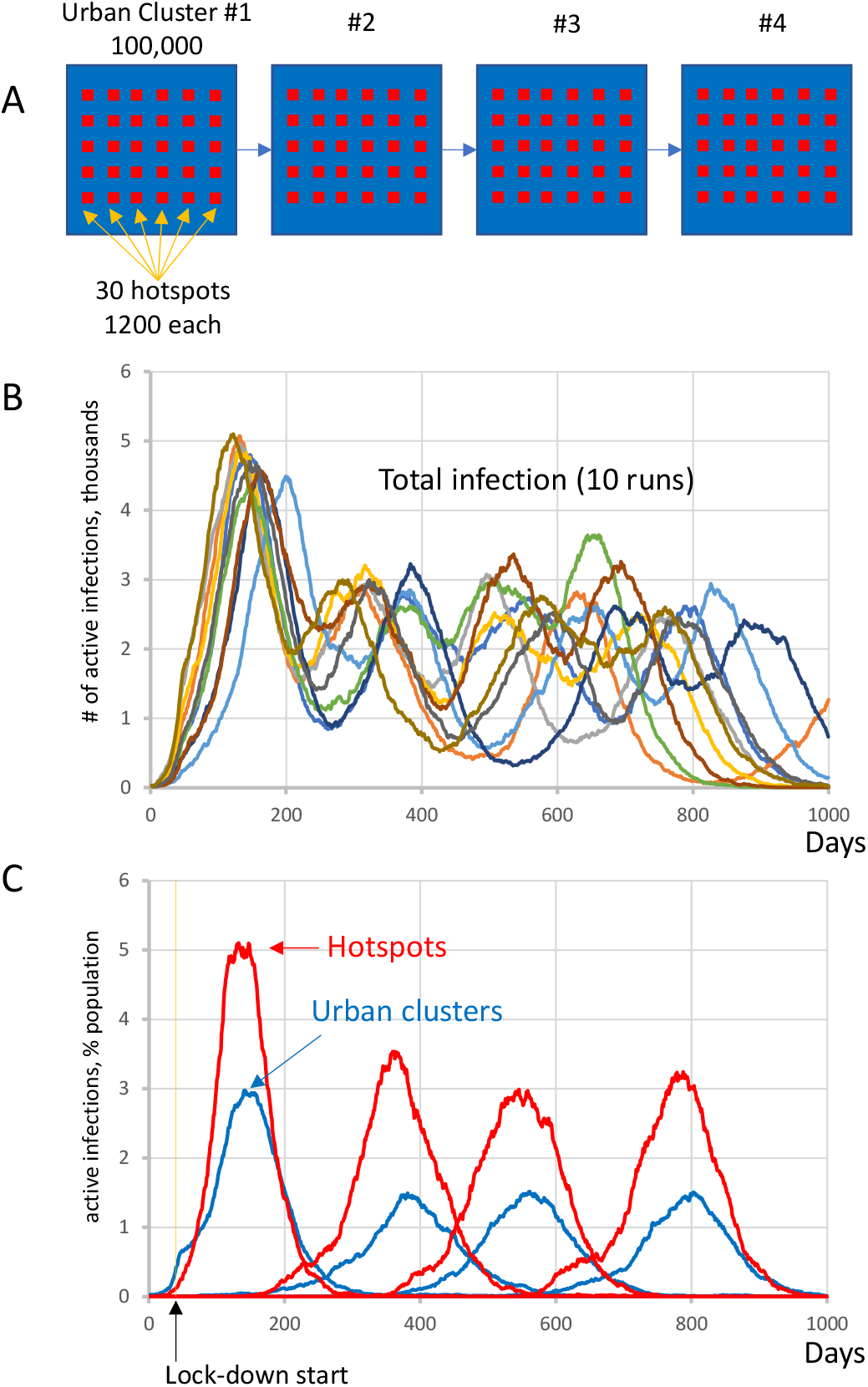
Complex infection propagation patterns in multiple urban areas containing hotspots. (**A)** Schematic illustration of the heterogeneous society used in simulations. (**B)** Total infection count oscillates as infection propagates. While individual oscillations exhibit substantial variations in timing and amplitude, the patterns remain the same (i.e. 4 oscillations, reflecting infection surge in each urban cluster). (**C)** The infection in hotspots is delayed before the lockdown at day 40, but then is always in the lead (red curves), driving infection in each urban cluster (blue curves) and facilitating infection propagation among clusters (Movie S2).

**Fig 4:**
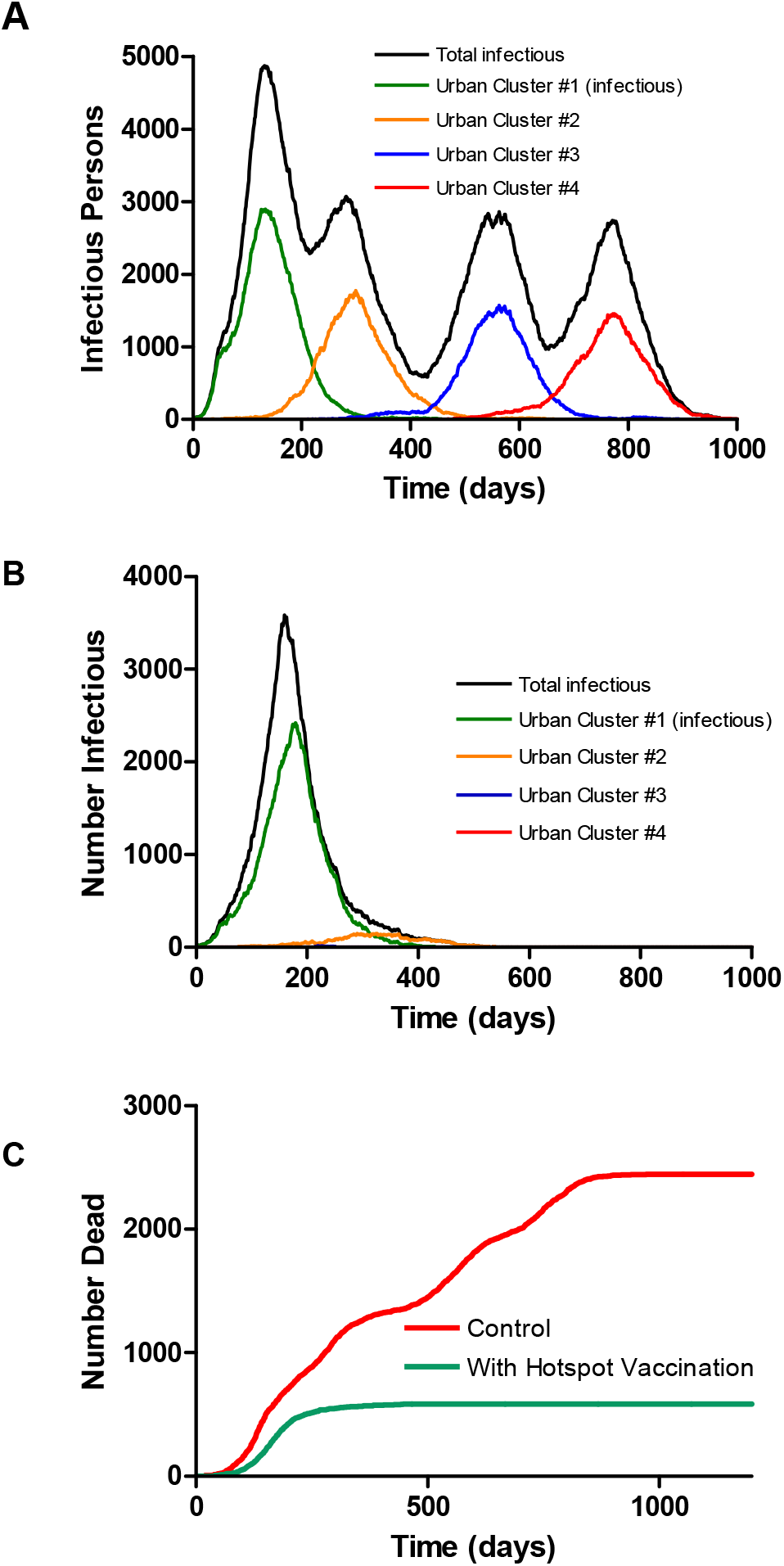
Multicity model as in figure 3 without (A) and with (B) vaccination of only the hotspot populations at a rate of 5% of the population per day, starting at day 150. Vaccination of hotspot individuals prevents geographic spread of the virus even though they are only about 1/3 of the population, thereby protecting the general population. The colored curves show only the infections among the socially distanced majority of the city population. (**C)** Overall mortality with and without vaccine, assuming case mortality of 1% in all groups.

### Reopening urban cluster after hotspots drive first wave of infection

We extended the single urban cluster hotspot scenario to reopen when infection numbers substantially drop. Here, the main cluster was composed of 1 million individuals which starts off closed with *R*_*0*_=1.05 and reopens to *R*_*0*_=2.50 at day 360. The cluster was connected to 30 potential hotspots each with 1200 +/- 500 people with *R*_*0*_=3.0 which remained constant throughout all simulation stages. The urban cluster was connected with 0.1% transient contact into the isolated clusters (*alphainpop*) while isolated clusters had 1% contact into the urban cluster (*alphaoutpop*). The results show two distinct waves of infection (Fig 5). The hotspots drove the first wave of infection, whereas the second wave was almost entirely composed of infection from the urban area, demonstrating that the hotspots acquired immunity and did not participate at all in the second wave. The ending of the first wave, dominated by the vulnerable groups, created the illusion that the epidemic was nearly over, while a large fraction of the surrounding populations was in fact still susceptible when reopening occurred.

**Fig 5:**
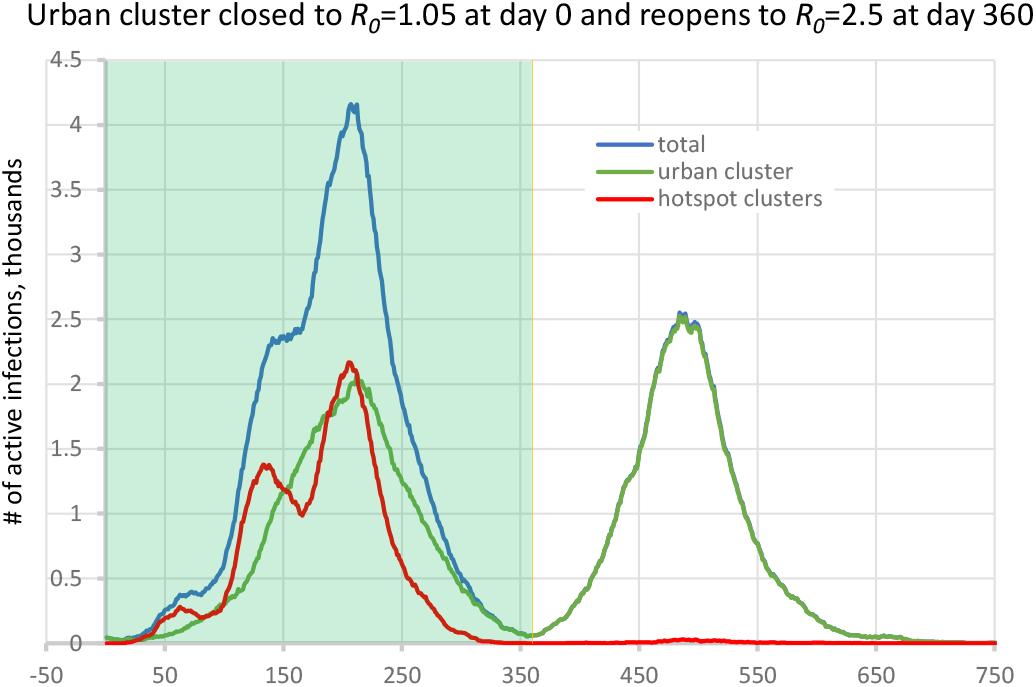
Second wave in the hotspot scenario. Urban cluster generates a second wave of infection when it reopens from *R*_*0*_=1.05 to *R*_*0*_=2.50 on day 360 (green line), whereas hotspots with *R*_*0*_=3.0 (red line) have acquired immunity in the first wave and do not participate in the second wave.

## Discussion

### Interpretations and implications

Since Summer of 2020, the infection curves of the COVID-19 pandemic in various locations have been very different from standard smooth bell curves. Here we tested the hypothesis that multiple, asynchronous waves and plateaus are in part due to stochasticity and heterogeneity, as well as due to changing efforts at mitigation. Geographic heterogeneity is included in forecasting models [12, 21, 22] which use extensive, public databases of population characteristics and travel patterns, but these do not fully account for the stratification of social behaviors that controls the spread of the virus. Therefore, instead of building another data-based forecasting and estimation model, we developed a numerical scenario model that we used to explore mechanisms of infection dynamics with regards to social stratification. The model was built as a network of “populations” which represent social and behavioral strata of geographic populations. Our model can be considered a metapopulation of SARS-CoV2, when a single species is spread among different environments that determine its local survival or extinction.

We examined several scenarios which included one or more large urban populations connected to vulnerable subgroups that are unable/unwilling to socially distance and thus represent potential COVID-19 hotspots. Depending on the degree of interaction, these subgroups were either driven by infection from the main population, or acted as major drivers of the epidemic. Isolated subpopulations were infection-driven (*e*.*g*. nursing homes, prisons, remote suburbs, clustered religious groups) and had a substantially delayed contribution to total infection cases, ultimately forming an infection curve which could include multi-modal growth periods, an extended plateau, a prolonged tail, or a delayed second wave of infection (Fig 1). These communities, due to their isolated nature, had low herd immunity that put them at risk for explosive scenarios when basic mitigation strategies were not implemented. Alternatively, partially integrated subpopulations were driving infection (*e*.*g*. employees of factories, warehouses, meat packing plants, church groups, campuses, shelters, and other essential workers) in its connected urban population by picking up infection and amplifying it by (Fig 2, movie S1). We found that these “hotspots” ignite infection even in a locked down population, ultimately propagating and igniting other isolated populations (Fig 3, movie S2). The locked down population however does not acquire herd immunity, as opposed to the hotspots, and thus when lockdown is lifted, a second wave is generated by the main cluster (Fig 5).

There are several implications that arise from our results. We can expect social heterogeneity to form delayed local asynchronous epidemics, creating a variety of infection profiles in various regions over time, prolonging the pandemic time span, and spreading to new areas unpredictably due to the stochasticity of infection in small subgroups, as is becoming increasingly obvious in the United States in the Fall of 2020 Effective mitigation of the epidemic in the main population requires close attention to vulnerable subgroups in order to prevent the formation of COVID-19 infection hotspots. Otherwise vulnerable subgroups that cannot implement mitigation strategies spread infection to the socially distanced populations, defeating their efforts at mitigation. Despite hotspots possibly acquiring immunity, there still exists a threat of a second wave of infection in the socially distanced main population. Thus, an effective treatment or vaccination needs to be developed prior to full reopening. As vaccines become readily available, the selection and timing of their administration will be an important policy consideration. Our simulations in idealized scenarios (Fig 4) suggest that focusing vaccination on the small fraction of the population that is unable or unwilling to socially distance may be sufficient to interrupt regional spread and protect a much wider fraction of the public. Notably, achieving this effect requires vaccinating all hotspot groups, not merely medical personnel, and essential workers, but also uncooperative college students and those with an aversion to mitigations. This creates a kind of moral hazard – rewarding bad behavior – but the model suggests that it is the public interest.

### Comparison with other studies

While our study is focused on vulnerable subpopulations in pandemic development, there are other important factors regarding social heterogeneity identified by previous studies. The study by Dolbeault et al. [23], using their multi-group SEIR model, underlined the importance of mitigation measures on single individuals with a high level of social interactions. Indeed, their study showed that even a small group of individuals with high transmission rate can trigger an outbreak even if the *R*_*0*_ of the majority is below 1. Althouse et al. [14] identified and explored in depth another important factor, explosive super-spreading events originating from long-term care facilities, prisons, meat-packing plants, fish factories, cruise ships, family gatherings, parties and night clubs. This study further demonstrated the urgent need for targeted interventions as routes of effective virus transmission. Taking into account the importance of these super-spreading events and individuals, they were included in the design of our model (see Methods, Super-spreading) to generate more realistic outcomes of scenarios.

With regard to agent-based models, Chinazzi et al. [12] used GLEaM to demonstrate that travel restrictions introduced in Wuhan in January 2020 only delayed epidemic progression by 3 to 5 days within China, and international travel restrictions only helped slow infectious spread until mid-February. Our simulations of COVID-19 spread also show that ultimately, when enough time goes by, isolation does not prevent infection of vulnerable subpopulations (Fig 1). Chinazzi et. al. suggests that early detection, hand washing, self-isolation, and household quarantine are more effective than travel restrictions at mitigating this COVID-19 pandemic. Our recommendations are in accord, and we advocate for communities to take extra care of vulnerable subpopulations internally, as so to prevent a possible hotspot formation that may evolve into a regional epidemic.

### Model features, limitations, and future studies

An epidemic can be likened to a forest fire, which spreads by diffusion along a front, but can also jump by embers that may or may not start a new blaze. Such spread to virgin areas, with a virus as with a fire, is intrinsically stochastic and such stochasticity, which is not explicitly included in mean-field models, may contribute to the remarkable patchiness of the COVID-19 epidemic. This has caused the epidemic to appear entirely different to observers in different locations, leading to politicization of the response, which is, itself, a form of social heterogeneity. For rare spread to small, isolated subgroups (embers) this stochasticity is crucial. Patchiness is aggravated by the over-dispersion (super-spreading) of secondary cases of COVID-19, where the majority of infected individuals do not spread the virus, but some can cause up to a hundred secondary infections [14]. Our model is explicitly stochastic, with a mechanism to account for over-dispersion, by keeping a partial history of individual infections. Furthermore, the design of our new model allows it to be applied in future studies of real-world scenarios on any scale, limited only by memory and the ability to determine the underlying topology and parameters.

However in our model, we make no attempt to distinguish between symptomatic and asymptomatic cases, despite recent findings by Chao et al. [24] in their agent-based model (dubbed Corvid) that demonstrated that most infections actually originate from pre-symptomatic people. Since the relative infectivity of symptomatic and non-symptomatic is uncertain, there is no direct way to accurately determine the number of asymptomatic infections at present. Such a distinction (included in a number of other models) could easily be added by subdividing the 5 compartments, at the risk of added complexity and more parameters needed in a scenario.

We did not take into account recent suggestions that infectivity is concentrated in a short time window just before and after symptom onset. Instead, we used the standard SEIRD assumption that infections are generated throughout the period of infection, using a mean clinical duration of 7 days. The model does not consider the physical mechanisms of transmission of COVID-19, or the possibility that many recovered patients do not quickly re-enter their normal social circles, delaying herd immunity. An additional compartment, with a pipeline mechanism, could also be added to account for this.

We examined several simple scenarios as a demonstration of our model, which revealed the important role of embedded, non-distancing sub-populations in infection propagation. Further studies require consideration of the role of model network topology. Several studies have shown that epidemic propagation in large, scale-free networks can result in the establishment of an endemic state even with small infection rates, preventing random vaccination from effectively ending the epidemic [25, 26]. Strictly speaking this cannot happen in the scenarios we considered, which assumed that recovered individuals are permanently immune – a choice we made because of the extreme rarity of re-infections with SARS-CoV-2. A more important point is that prior theoretical analyses pertained to networks of individuals, each of whom can be either infected or susceptible. Within a single population cluster, over-dispersed link distributions such as in scale-free networks can enable persistence of infection because highly connected individuals can scavenge rare infections and widely redistribute them [27]. This is a major mechanism of super-spreading, which is incorporated in our model by heuristically handling super-spreading in each homogeneous cluster. However, stratification of the connectivity of individuals is not included in the model: Individual villages were taken to be homogeneous, characterized by their populations, *R*_*0*_ and r_eff_ that determine the effective dispersion of secondary infections. Further stratification of individual connectivity could be handled by splitting social behavior into separate, mutually embedded clusters e.g. college students who study together vs. those who study alone

It requires further studies to see if similar topological considerations pertain to networks of populations as in our model. With that in mind, the model includes the possibility that a recovered individual may revert to being susceptible, with a specified rate constant. How the topology of the larger-scale network of populations affects the propagation of the virus requires simulation studies too extensive to be considered in this paper. For example, whether physical transportation and communication networks are scale-free is controversial [28-30]. In our preliminary simulations (not shown), we found that a scale-free random network of 500 villages with populations proportional to the link numbers, and uniform behavior, had a significant probability of entering an endemic state even when the lifetime of immunity was as long as 500 days. However, the same was true of Erdös-Rényi random networks with a similar number of links. Interestingly, both types of random networks produced smooth single-peak epidemics resembling a single population suggesting that the increasingly complex patterns now being observed do depend on behavioral heterogeneity.

## Supporting information

Model code and parameters

## Data Availability

All data is available in the manuscript and supplementary materials.

## Data Availability Statement

The raw data supporting the conclusions of this article will be made available by the authors, without undue reservation.

## Author Contributions

All authors listed have made a substantial, direct and intellectual contribution to the work, and approved it for publication.

## Acknowledgments

This manuscript has been released as a pre-print at https://www.medrxiv.org/content/10.1101/2020.07.10.20150813v3 [31]. This study utilized the high-performance computational capabilities of the Biowulf Linux cluster at the National Institutes of Health, Bethesda, MD. (https://hpc.nih.gov/)

## Supplementary Material for

**Fig. S1.**
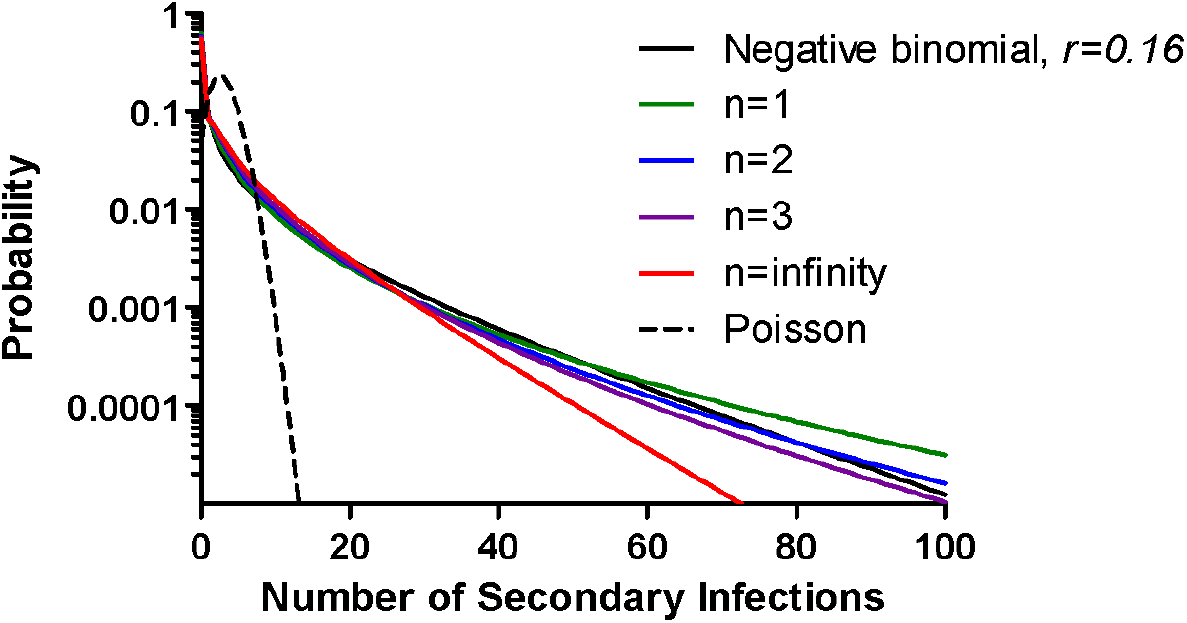
The distribution of secondary infections generated by infectious individuals. Black: Observed negative binomial distribution (Althouse, B. M., et al. 2020; “Stochasticity and heterogeneity in the transmission dynamics of SARS-CoV-2.” https://arxiv.org/abs/2005.13689); Green, blue, magenta, red: The actual realized number of secondary cases generated over the lifetime of one infection in the presence of n other infections individuals according to our scheme. All distributions have mean R0 =3.0. Dashed line: Poisson distribution with mean R0 as implicit in mean-field SEIR models.

**Fig. S2.**
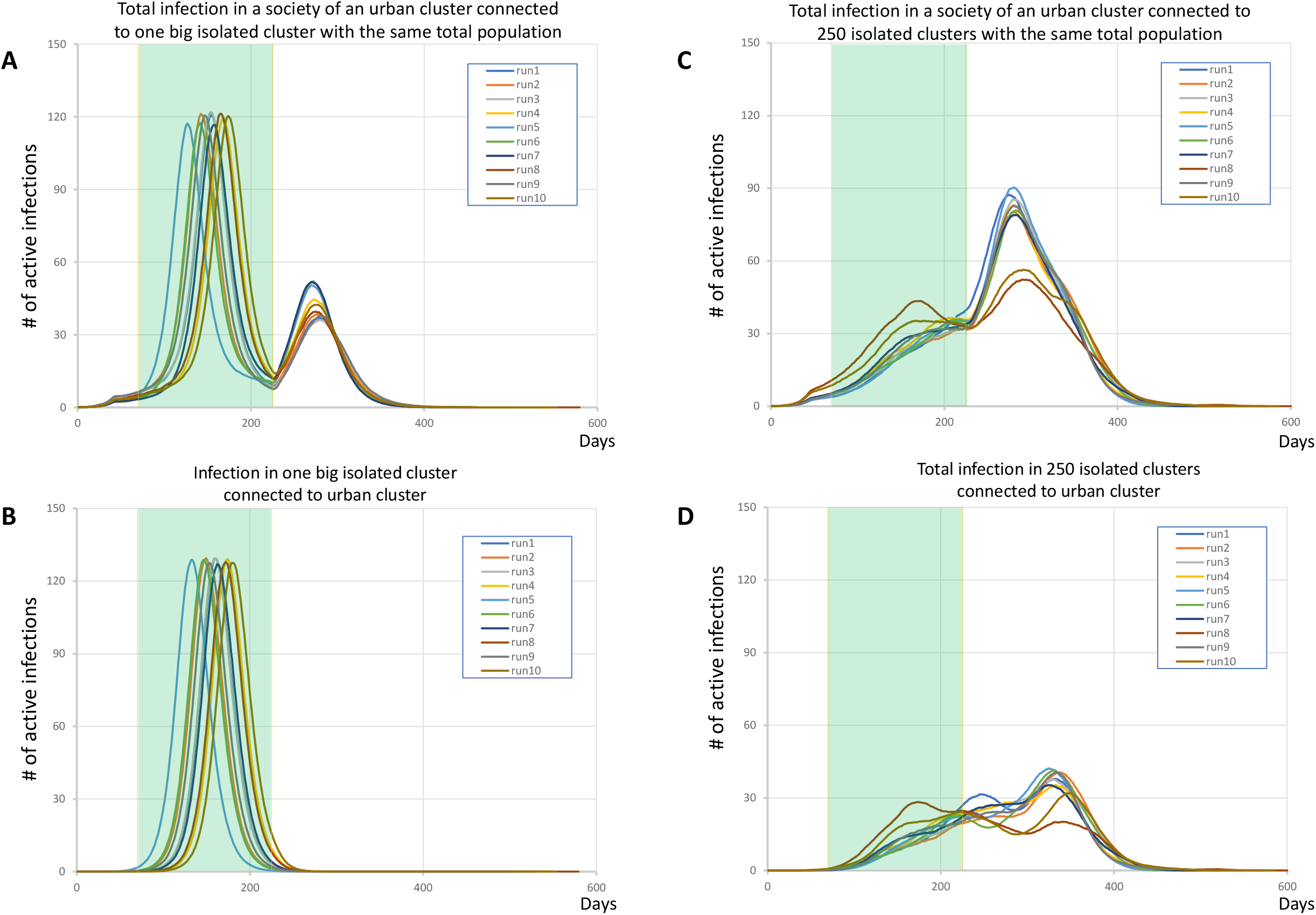
Heterogeneity of isolated clusters is important for the infection pattern. In the most complex scenario of “moderate lockdown” (Fig. 1C in main text) we substituted 250 clusters by one big cluster with the same population of one million people keeping all other parameters the same. The big isolated cluster generated substantial and sharp infection peak (**panels A and B**) that is absent or very small in case of 250 isolated clusters (**panels C and D**). Each panel shows 10 simulation runs (overlapped multi-color curves). The lockdown period from day 40 to day 225 is shown by green shade.

**Fig. S3.**
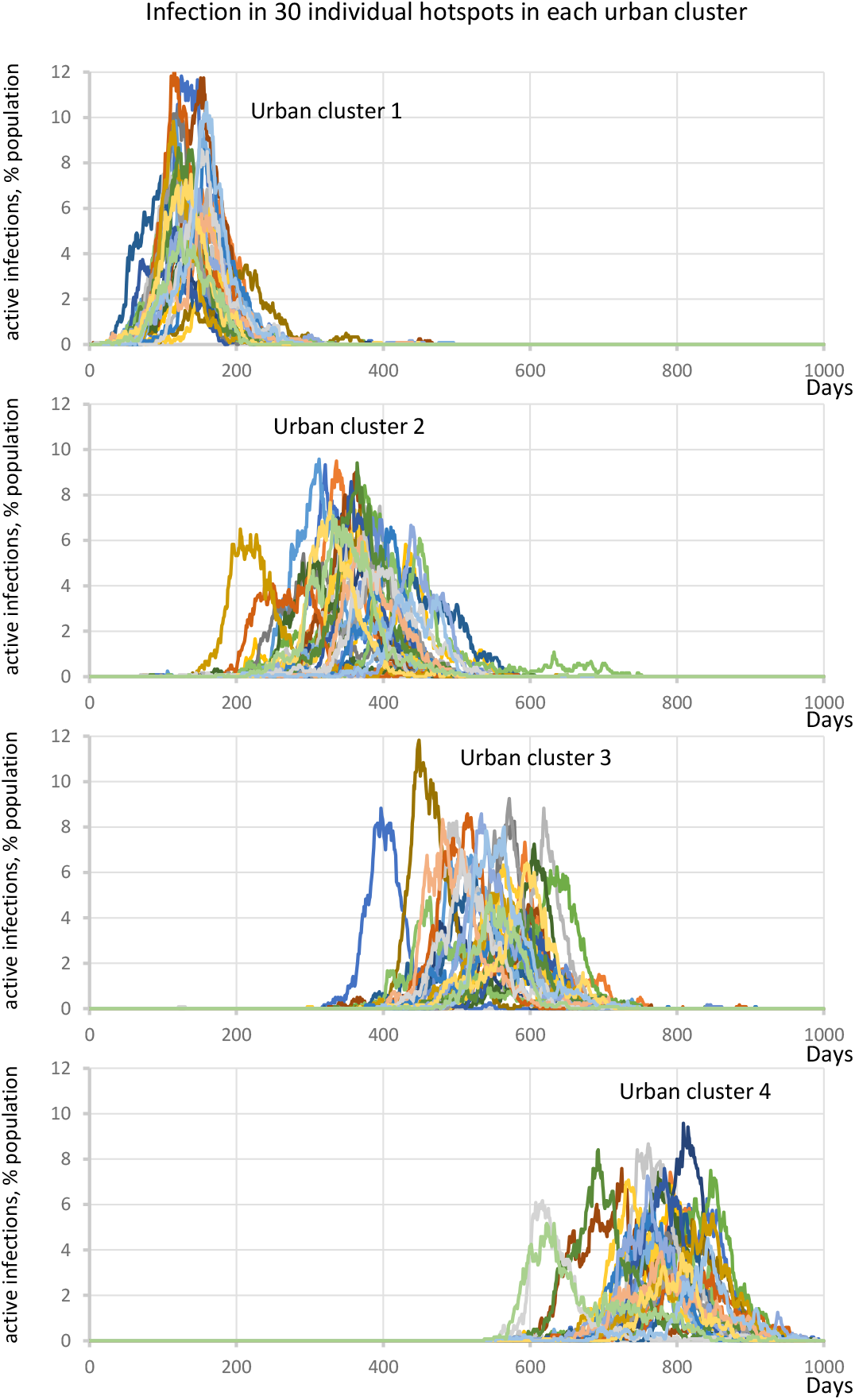
Stochastic propagation of infection from one urban area to another via hotspots in a society of 4 connected urban areas, each in lockdown but having hotspots. Each plot from top to bottom shows infection explosions in each individual hotspot for each urban area (specified by labels). See main text for details and also Movie S2.

**Movie S1 (separate file). Highly susceptible integrated clusters (hotspots) drive SARS-Cov2 infection in an urban cluster in stochastic simulations of SHEM model**. Infection time-dependent changes in hotspots (small squares) are coded by red shades saturating (pure red) at 5% of infection in each individual cluster. Infection in the urban area (big square) are is coded by blue shades saturating (pure blue) at 5% of infection in the area. The time is shown in the left upper corner in number of days. Large urban cluster had 1 million individuals with *R*_*0*_ = 1.25 while 250 hotspot clusters with 1200 +/- 500 people had the same internal *R*_*0*_ = 3.0.

**Movie S2 (separate file). Hotspots drive SARS-Cov2 infection in each urban cluster and infection propagation among urban clusters in stochastic simulations of SHEM model in a society of 4 connected urban areas**. Time-dependent changes of infection in hotspots (small squares) are coded by red shades saturating (pure red) at 3% of infection in each individual cluster. Infection in the urban area (big square) is coded by blue shades saturating (pure blue) at 3% of infection in the area. The time is shown in the left upper corner in number of days. Each urban area of 100,000 people at day 40 became closed from *R*_*0*_ = 2.5 to *R*_*0*_ = 1.25 at day 40. Each urban area has 30 hotspots with 1200 +/- 500 people that avoid closing and keep the same internal *R*_*0*_ = 2.

