## Supplementary material for "Social heterogeneity drives complex patterns of the COVID-19 pandemic: insights from a novel Stochastic Heterogeneous Epidemic Model (SHEM)": Model code and parameters: README.txt

SYSTEM: Linux
This README explains how to execute an epidemic simulation using SHEM 
and its corresponding parameters, using one of the demonstration scenarios presented in the paper.

For user creation of more general scenarios, the simulation engine module SIMULATOR contained in
the file simmod.for must be compiled first. The user then creates a front-end program that sets
up the topology and parameters of the simulation.  The front end then calls the subroutine episim
contained in the module, repeatedly for each time period (e.g. open, lockdown, reopen, etc.).
The parameters rinn (array indexed by populations), alphain, alphaout, betain, betaout (arrays
indexed by links) may be changed between calls.  The logical flag yflag should be queried to determine
when the simulation has finally concluded and the results extracted from the arrays tstep and kout.
Full details of the calling sequence are included in the comments in simmod16.for.

First step is to install gfortran:
sudo apt-get install gfortran

Using gfortran we compile the code as such in this order:

Single Thread:
gfortran simmod16.for epichain16F.for

Parallel:
gfortran simmod16_parallel.for epichain16F.for -qopenmp

Parallel only useful if 8 or more threads available
Performance on multithreaded Linux may be improved by: 
export KMP_AFFINITY=compact

An executable named "a.out" should appear.
Rename the executable to "SHEM" using:
mv a.out SHEM

Next we create a parameter file with the name "epistoch.dat"
Example parameters are supplied in this zip file (make sure to rename them to epistoch.dat).
Parameter descriptions are at the end of this README.
Output file decription is also at the end of this README.

To execute the program run:
swnum=1 filenum=1 ./SHEM

The program can be run multiple times in parallel by changing filenum
to a different number and executing the program again. swnum should
be set to the largest number of parallel realizations being scripted.

======================
PARAMETER DESCRIPTIONS
======================

General Parameters
pname = embedded name of results
tbin = time step for output purposes
nh = total cities connected serially
tswitch1 = start of the closed period
tswitch = start of the reopen period
spreads= if true (1) then a visitor keeps his superspreading capabilities from the previous enviornment,
	if false (-1) then a visitor gets new superspreading capabilities from the new enviornment

Subpopulation 1: Urban Cluster
r00 = R0 of the urban cluster during the open stage
r01 = R0 of the urban cluster during the closed stage
r02 = R0 of the urban cluster during the reopen stage
kstot = population of the urban cluster
i0 = the integer number of infectious individuals at initialization in the urban cluster
kx0 = the integer number of exposed individuals at initialization in the urban cluster
amu0 = mortality rate per infection for the urban cluster (constant)
btinpop = rate of people moving into adjacent urban cluters (open + closed)
btoutpop = rate of people moving out of adjacent urban cluters (open + closed)
btinpop2 = rate of people moving into adjacent urban cluters (reopen)
btoutpop2 = rate of people moving out adjacent urban cluters (reopen)
vax0 = the fraction of the urban cluster that is vaccinated per day starting at tswitch

Subpopulation 2: Isolated/Integrated Clusters
amu2 = mortality rate per infection for subpopulation 2 (constant)
alphinpop = the fraction of the urban cluster that visits a subpopulation 2 cluster on average (open + closed)
alphinpop2 = the fraction of the urban cluster that visits a subpopulation 2 cluster on average (reopen)
alphoutpop = the fraction of a subpopulation 2 cluster that visits the urban cluster on average (open + closed)
alphoutpop2 = the fraction of a subpopulation 2 cluster that visits the urban cluster on average (reopen)
anbar = expected population of each subpopulation 2 cluster
anvar = population variability of each subpopulation 2 cluster
rinpop = internal R0 of subpopulation 2 (open + closed)
rinpop2 = internal R0 of subpopulation 2 (reopen)
npop = total number of subpopulation 2 clusters
vaxpop = the fraction of the subpopulation clusters that are vaccinated per day starting at tswitch

========================
OUTPUT FILE DESCRIPTIONS
========================
The output file contains the SEIRD statistics of each subpopulation cluster at every time step of the simulation.
Note: SEIRD - Susceptible, Exposed, Infected, Recovered, Dead

Column 1 
Time

Column 2:6 
SEIRD totals in all populations (in that order, each state has its own column).

Column 7:7+nh*([1+npop]*5)-1
SEIRD is printed sequentially for each urban cluster and its corresponding subclusters.
For example, the output format for nh=2, npop=3 is: [A B B B A B B B] where A is the SEIRD of the urban 
cluster and B is the SEIRD of its corresponding subclusters
